# Cervical Cancer Screening in South Florida Veteran Population 2014-2020: Cytology and High-Risk HPV Correlation and HPV Epidemiology

**DOI:** 10.1101/2021.06.10.21258483

**Authors:** Lee B. Syler, Corinne L. Stobaugh, Philip R. Foulis, George T. Carlton, Lauren A. DeLand, Andrew A. Borkowski

**Affiliations:** University of South Florida; James A. Haley Veterans’ Hospital; James A Haley Veterans’ Hospital

## Abstract

**Objective:** This project aims to use our robust women’s health patient data to analyze the correlation between cytology and Hr-HPV testing, study performance of Hr-HPV testing for detecting cytology lesions, and examine epidemiologic measures of HPV infections in the women’s veteran population.

**Methods:** We collected patient data from 2014 to 2020 from our computerized patient record system. We performed HPV assays using the ROCHE 4800 system. The COBAS HPV assay detects HPV 16, HPV 18, and 12 other HPV types (31, 33, 35, 39, 45, 51, 56, 58, 59, 66, and 68). We organized cytology results and Hr-HPV assays with Microsoft Access and Microsoft Excel for analysis.

**Results:** A total of 9437 cervical specimens were co-tested. High-grade cytology lesions (HSIL or higher and ASC-H) were overwhelmingly positive for Hr-HPV (94.1% and 87.2%, respectively). Low-grade cytology lesions (LSIL and ASC-US) were positive for Hr-HPV in lower percentages (72.6% and 54.9%, respectively). Hr-HPV testing had a sensitivity of 91.3%, a specificity of 93.1%, a positive predictive value of 16.4%, and a negative predictive value of 99.8% for detecting high-grade cytology lesions. Hr-HPV testing had a lower performance for detecting low-grade cytology lesions. Ten (10) cases had high-grade cytology and negative Hr-HPV test. Nine out of ten (9/10) of these patients showed no dysplasia (6) or low-grade dysplasia (3) on subsequent biopsy. Overall, 14.4% of tests were positive for Hr-HPV. The highest positive Hr-HPV test rates were in the second and eighth decades of life, 25.1% and 22.0%, respectively. In women over age 30, HPV types 16 and 18 were present in 11.7% and 6.4% of tests, respectively. Other HPV types were present in 82.3% of tests.

**Conclusions:** Hr-HPV testing has high performance for detecting high-grade cytology lesions. We believe our findings are in accordance with recent studies and guidelines that recommend primary Hr-HPV testing as the preferred screening method. The percentage of positive Hr-HPV tests and rates for age and HPV types 16 and 18 in our women’s veteran population suggest similar HPV prevalence to that of the general US population.

## Introduction

Human papillomavirus (HPV) is one of the most potent carcinogens in humans, with almost 5% of all new cancers diagnosed worldwide attributable to HPV [1]. These cancers include cervical, anal, vaginal, penile, vulvar, and oropharyngeal. Virtually 100% of cervical cancers are HPV-related [1]. HPV is estimated to be the most common sexually transmitted disease infection (STD) in the United States [2]. Although broad spread screening practices and vaccination have significantly decreased the incidence and mortality of cervical cancer in the US, an estimated 14,480 new cervical cancer diagnoses and 4,290 deaths will occur in 2021 [3].

HPV infects epithelial cells, promoting cellular proliferation, blocking apoptosis, and evading the immune system [4-7]. Persistent infection is necessary for the development of cervical intraepithelial neoplasia (CIN), and the probability of complete clearance is dependent on the duration of infection [8, 9]. Important factors that determine infection progression are the HPV genotype and immunosuppression [10]. Their carcinogenic effects have classified HPV genotypes as high-risk HPV’s (Hr-HPV) and low-risk HPV’s (Lr-HPV). Of the Hr-HPV types, types 16 and 18 are the most persistent and carcinogenic [11, 12]. Thankfully, the vast majority of HPV infections, roughly 90%, are cleared by the immune system within the first year despite immune evasion mechanisms of the virus [11].

Since the 1950s, for most of this period, the basis of cervical cancer screening has relied on the revolutionary Papanicolaou test and, later, liquid-based cytology. As cervical screening technologies evolved and knowledge of the natural history of cervical cancer grew, so did the recommendations for screening and management.

One of the most substantial changes in screening and management guidelines was the incorporation of Hr-HPV molecular testing into screening protocols for cervical cancer by the American Society for Colposcopy and Cervical Pathology (ASCCP) in 2012 [13]. At that time, they did not recommend primary Hr-HPV testing (only Hr-HPV testing without cytology) for screening due to concerns of specificity and possible excess treatment of non-neoplastic HPV lesions.

In 2015, after growing evidence of the high performance of primary Hr-HPV testing [14-21] and the FDA approval of a Hr-HPV assay for primary Hr-HPV screening, the ASCCP published an Interim Clinical Guidance for the use of primary Hr-HPV testing for cervical cancer screening [22].

In 2020, the American Cancer Society (ASC) updated the cervical cancer screening guidelines. They recommended primary HPV testing as the preferred screening method, with co-testing and cytology alone being acceptable [23]. They also mentioned that the guidelines should be transitional towards primary HPV testing and that co-testing and cytology alone should gradually phase out. The current FDA-approved HPV assays for primary HPV screening are Cobas HPV and Onclarity HPV. The FDA approves these and other assays for co-testing [23].

During the last decades, the Veterans Health Administration (VHA) has seen a significant increase in its women patient population. The growing numbers of women participating in the US military have made this subgroup the fastest growing subgroup of US veterans [24]. The VHA commits to delivering comprehensive primary care for women. A Designated Women’s Health Primary Care Provider (DW-PCP), who leads a Women’s Health Patient Align Care Team (WH-PACT), provides this care. The DW-PCP and WH-PACT must have sufficient training and expertise to care for women veterans [25].

Although recent cervical screening guidelines shift towards primary HPV screening as the main method for cervical cancer screening, we use co-testing at James A. Haley Veterans Affairs Hospital. Co-testing provides an excellent opportunity for comparing and correlating cytology and Hr-HPV testing results. Studies that compare cytology testing to Hr-HPV testing in the US women’s veteran population are lacking. As the gradual shift towards primary Hr-HPV screening occurs, we believe more studies will compile in favor of the performance of Hr-HPV testing in different scenarios.

Female veterans have a higher prevalence of interpersonal trauma due to their military service, affecting their healthcare needs [26-28]. Although it is not yet clear if veteran women have a higher prevalence of HPV infection and cervical cancer than the general population, some studies suggest that this may be the case [29-31]. Although researchers have performed studies on HPV prevalence in the general population of the US [32-34], these studies are lacking in the US women veteran population. More studies on cervical cancer and HPV prevalence affecting the US women’s veteran population are needed.

This project aims to use our robust women’s veteran health data to analyze the correlation between Hr-HPV testing and cervical cytology. In doing so, we measure the performance of Hr-HPV testing for the detection of high-grade versus low-grade lesions diagnosed by cytology within this specific population. We also calculate the percent of positive Hr-HPV tests among US veteran women, a population in which these studies are scant.

## Methods

A total of 9437 cervical specimens of women veterans were co-tested by Hr-HPV testing and cytology from 2014 through 2020.

Hr-HPV testing was done through the COBAS HPV assay using the ROCHE 4800 system. The COBAS HPV assay detects HPV 16, HPV 18, and 12 other HPV types (31, 33, 35, 39, 45, 51, 52, 56, 58, 59, 66, and 68). Two major processes are the basis of the assay: (1) automated specimen preparation to simultaneously extract HPV and cellular DNA; (2) PCR amplification of target DNA sequences using both HPV and B-globin specific complementary primer pairs and real-time detection of cleaved fluorescent-labeled HPV and B-globin specific oligonucleotide detection probes. The concurrent extraction, amplification, and detection of B-globin in the Cobas HPV Test monitor the entire test process.

The master mix reagent for the Cobas HPV Test contains primer pairs and probes specific for the 14 high-risk HPV types and B-globin DNA. The amplified DNA (amplicon) detection is performed during thermal cycling using oligonucleotide probes labeled with four different fluorescent dyes. The amplified signal from 12 high-risk HPV types (31, 33, 35, 39, 45, 51, 52, 56, 58, 59, 66, and 68) is detected using the same fluorescent dye. In contrast, specific fluorescent dyes detect HPV 16, HPV 18, and B-globin signals.

We divided cytology results into two main diagnostic groups: high-grade cytology lesions, including ASC-H and HSIL or higher, and low-grade cytology lesions, including ASC-US and LSIL. To measure the correlation between Hr-HPV testing and cytology and to measure Hr-HPV testing performance for detecting positive cytology lesions, results of cytology and Hr-HPV assays were sorted, organized, and analyzed using Microsoft Access and Microsoft Excel.

We calculated percentages of Hr-HPV positive cases for high-grade and low-grade cytology lesions and sensitivity, specificity, positive predictive value, and negative predictive value of Hr-HPV testing for detecting high-grade and low-grade cytology lesions. We further analyzed cases that showed high-grade cytology but resulted in a negative Hr-HPV test looking for common findings and possible causes of discrepancy. Cases showing negative cytology that resulted in a positive Hr-HPV test were not further analyzed since not all Hr-HPV infections develop high-grade dysplasia.

We calculated the total percentage of Hr-HPV positive cases. We divided women by age groups (20-29, 30-39, 40-49, 50-59, 60-69, 70-79, and >80) and calculated the percentages of positive Hr-HPV in each age group. In women over age 30, we determine the rate of positive tests for HPV types 16, 18, and other types precisely. The reason for leaving out women under age 30 is that we only report Hr-HPV type results in women over age 30 with a cytology result of ASC-US, LSIL, or higher.

## Results

A total of 9437 cervical specimens of women veterans were co-tested during the 2014-2020 period. The Hr-HPV test results for high-grade cytology lesions (HSIL or higher and ASC-H) were overwhelmingly positive for Hr-HPV (94.1% and 87.2%, respectively) *(Chart 1)*.

**CHART 1.**
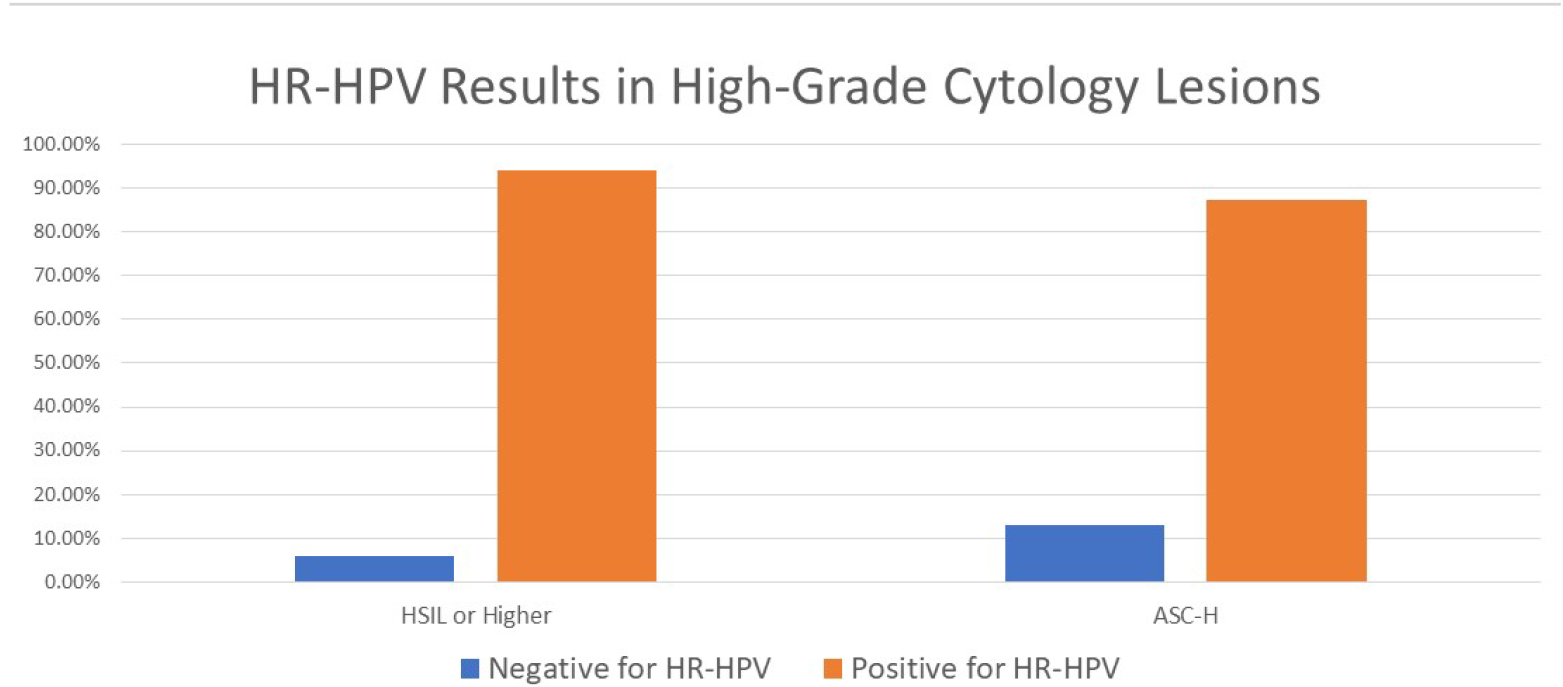

Low-grade cytology lesions (LSIL and ASC-US) were positive for HR-HPV in lower percentages (72.6% and 54.9%, respectively) *(Chart 2)*.

**CHART 2.**
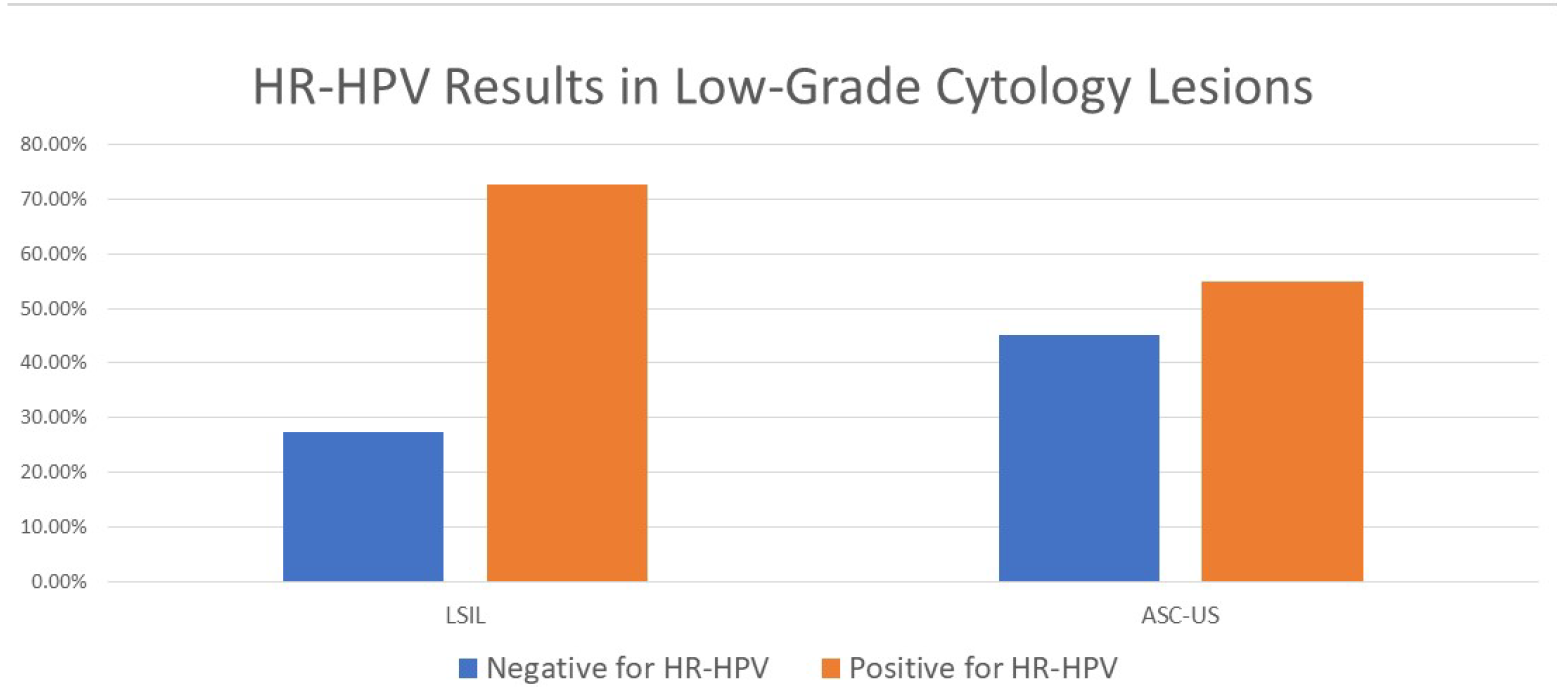

Hr-HPV testing had a sensitivity of 91.3%, a specificity of 93.1%, a positive predictive value (PPV) of 16.4%, and a negative predictive value (NPV) of 99.8% for detecting high-grade cytology lesions. However, it had a lower sensitivity of 58.9%, a specificity of 93.1%, a PPV of 56.2%, and a NPV of 93.8% for detecting low-grade cytology lesions. *Chart 3* compares these validity measures of Hr-HPV testing for high-grade versus low-grade lesions diagnosed by cytology.

**CHART 3.**
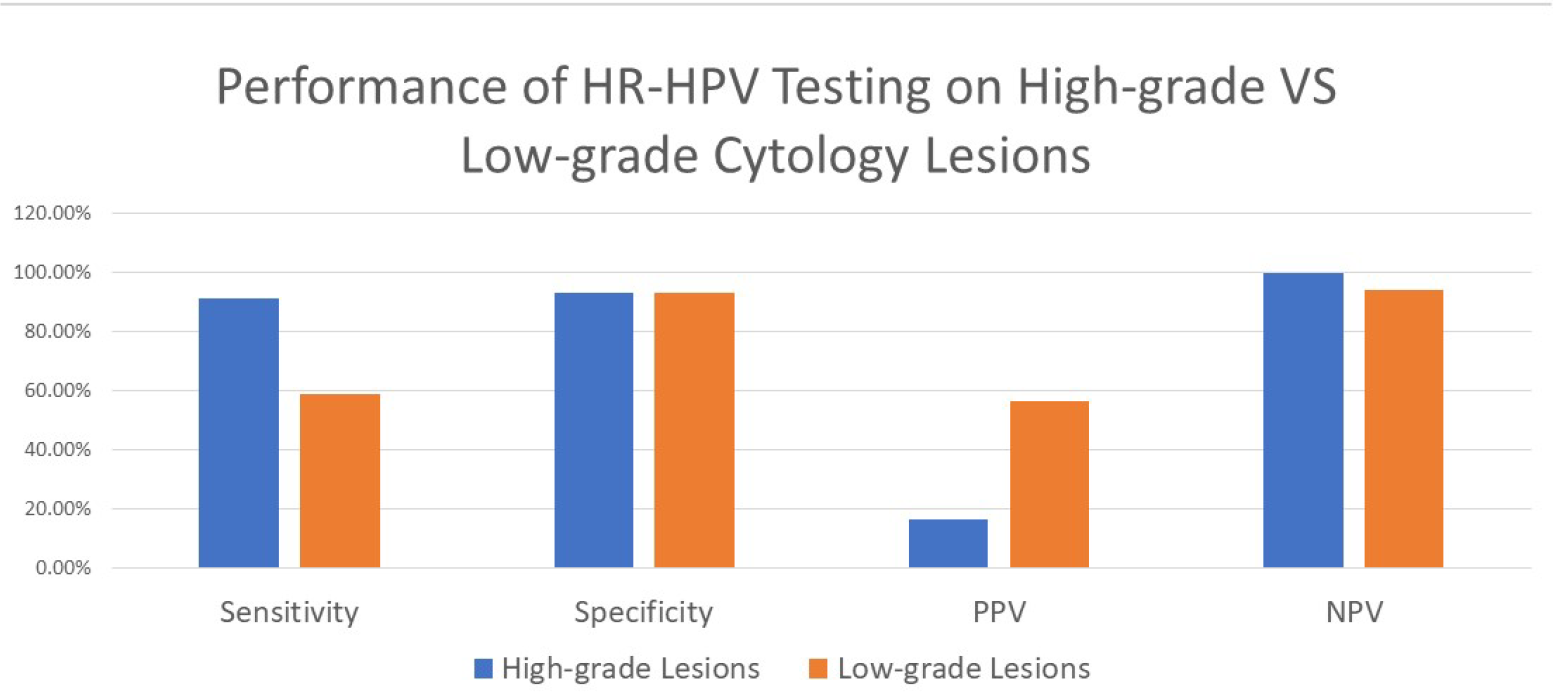

A review of medical records of patients with high-risk lesions on cytology and negative Hr-HPV showed that nine out of ten (9/10) had either no dysplasia (6) or low-grade dysplasia (3) on subsequent biopsy. Two of these cases showed microglandular hyperplasia; one of these simultaneously with low-grade dysplasia. The remaining case was diagnosed cytologically as ASC-H but was lost to follow-up and did not undergo biopsy. Cytology review of most of these cases identified significant background inflammation complicating cytologic interpretation.

Out of the 9437 cervical specimens, 1359 cases were positive for Hr-HPV, constituting 14.4% *(Table 1)*.

**TABLE 1.**
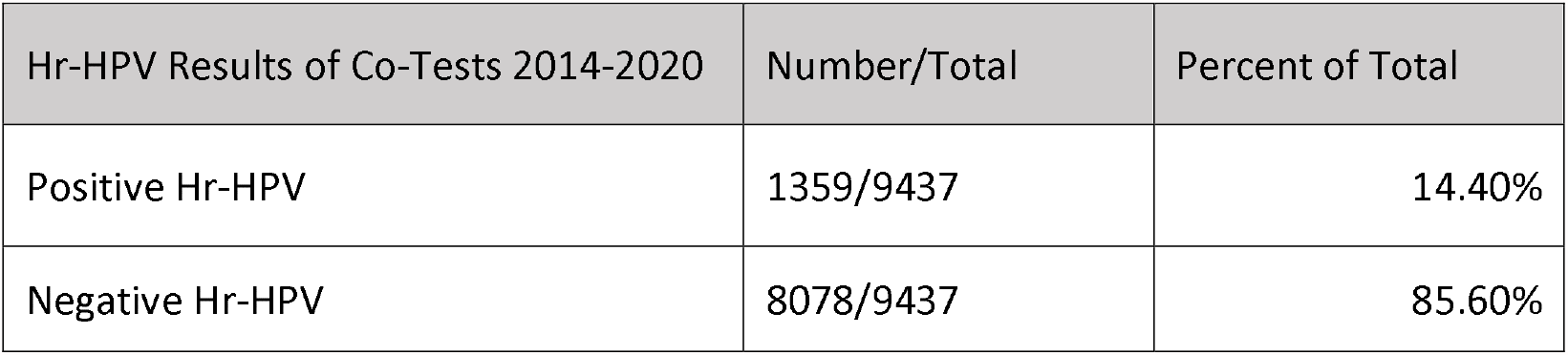

Description of age group-specific positive Hr-HPV percentages is in *Table 2*. The highest positive Hr-HPV test rates were in the third and eighth decades of life, 25% and 22%, respectively.

**TABLE 2.**

| Age Group | Total Number | Number of Positive Hr-HPV | Percent Positive |
| --- | --- | --- | --- |
| 20-29 | 1325 | 333 | 25.1% |
| 30-39 | 2639 | 440 | 16.7% |
| 40-49 | 2274 | 254 | 11.2% |
| 50-59 | 2127 | 223 | 10.5% |
| 60-69 | 1015 | 98 | 9.7% |
| 70-79 | 50 | 11 | 22.0% |
| >80 | 7 | 0 | 0.0% |
| Total | 9437 | 1359 | 14.4% |

In women over age 30, 1036 were positive for Hr-HPV. Of these cases, 121 contained HPV type 16 as a single or coinfection (11.7%), 66 contained HPV type 18 (6.4%) as a single or coinfection, and 852 contained Hr-HPV types other than 16 and 18 (82.3%) *(Table 3)*.

**TABLE 3.**

| Hr-HPV Infection Type in Women Over Age 30 | Number/Total | Percent of Total |
| --- | --- | --- |
| HPV type 16 as single or coinfection | 121/1036 | 11.70% |
| HPV type 18 as single or coinfection | 66/1036 | 6.40% |
| Hr-HPV types other than 16 and 18 | 852/1036 | 82.30% |

## Discussion

Some of the most extensive studies worldwide have demonstrated the superior performance of primary Hr-HPV screening compared to cytology screening alone [18-21]. These studies have shown that incidence risks for cervical intraepithelial neoplasia grade 3 and higher (CIN3+) are higher in women screened with cytology alone when compared to those screened with primary Hr-HPV testing. But what about co-testing? Wouldn’t the combination of both screening strategies add to higher performance? Gage et al. answered this question in their study of close to 1 million screened women. They showed that the 3-year risk for developing CIN3 and cancer after a negative Hr-HPV test was lower when compared to the 5-year risk for developing CIN3 and cancer after a negative co-test. This finding suggested that the co-test derived most of its reassurance from the Hr-HPV testing portion.

In their study results from the ATHENA study, Write et al. were also able to demonstrate an increased risk of CIN3+ in women screened with cytology alone than those screened by Hr-HPV testing. They showed that Hr-HPV testing had a sensitivity of 76.1% for detecting CIN3+, higher than the 47.8% and 61.7% for cytology and co-testing, respectively.

Although we did not follow our patients to calculate incidence risks of CIN3 or cancer in our population, our results show a high positive correlation between high-grade cytology lesions and positive Hr-HPV testing results. The majority of high-grade cytology lesions were positive for Hr-HPV (94.1% of HSIL and higher, 87.2% for ASC-H). Hr-HPV testing was highly sensitive and specific for detecting high-grade cytology lesions, 91.3%, and 93.1%, respectively. It also had a near-perfect NPV of 99.8%, which gives high reassurance to those women with a negative Hr-HPV result. A low PPV of 16.4% is obvious since HPV types 16 and 18, together, have a 3-year cumulative incidence risk (CIR) of only 21.16% for developing CIN3+, and the other Hr-HPV’s have a 3-year CIR of 5.4% for developing CIN3+, according to the ATHENA study [20]. This means that not all Hr-HPV infections will progress to high-grade dysplasia, hence the low PPV.

When reviewing positive high-grade cytology lesions that were negative for Hr-HPV testing, we found that the majority had significant background inflammation complicating cytologic interpretation. On subsequent biopsy, nine out of ten (9/10) were either negative for dysplasia (6) or had low-grade dysplasia (3). Two of the revised biopsies showed microglandular hyperplasia, one of them with simultaneous low-grade dysplasia. Microglandular hyperplasia is a benign alteration of endocervical epithelium in which the endocervical cells can show reactive changes. These changes can be confused with a wide range of differential diagnoses during cytological interpretation, including LSIL, HSIL, AIS, and invasive cancer [35]. These are only some examples of situations in which cytological interpretation can be equivocal.

A concerning finding in our study is the low sensitivity of Hr-HPV testing for detecting low-grade cytology lesions. One cannot help but ask: What is the impact of missing low-grade lesions? Low-grade squamous intraepithelial lesions (LSIL), diagnosed by cytology, can be caused by Hr-HPV types or Lr-HPV types. A study from The Netherlands showed that, over four years, all women with LSIL, infected with low-risk HPV types, regressed to normal cytology, as did 70% of those infected with high-risk HPV types [36]. Another study done in Brazil showed that more than 90% of women with a cytology result of LSIL regressed within 24 months [37]. These studies suggest that not detecting a percent of low-grade cytology lesions through primary Hr-HPV testing would not significantly impact screening for cervical cancer, especially since Lr-HPV types would cause the missed cases.

Our results show that 14.4% of all Hr-HPV tests were positive in our women’s veteran population. Our literature search for the overall prevalence of Hr-HPV types in the US general population found variable results. A study carried out by Monsonego et al. used extensive data from the ATHENA trial with over 40,000 women screened in 23 states. They showed the lowest overall Hr-HPV prevalence, 13.4% [34], which is very close to our calculations of the total percent of positive Hr-HPV tests. Another study showed a similar prevalence of 15.2% [32], and another showed a higher prevalence of 20.4% [33] among the general US female population. These numbers suggest that, in the women’s veteran population that we serve, Hr-HPV prevalence is not higher than that of the general US population; in fact, it suggests similarity.

When analyzing age-specific rates of Hr-HPV positive tests, we found them very similar to the rates of the general women’s public calculated by Wright et al. [20] from the ATHENA study results (See *Table 4*). Our 20-29 age group had the highest Hr-HPV positive rate, similar to their 25-29 age group, 25% versus 21%, respectively. Our data show a second spike in positive rates in the eighth decade of life (22%). Wright et al. did not further subdivide their age groups past 50; hence we could not compare to see if a similar spike exists in the general population. However, our number of cases is small in this age group; only 50 patients.

**TABLE 4.**

| Our US Women's Veteran Population |  | General US Population From ATHENA Study Results (Wright et al.) |  |
| --- | --- | --- | --- |
| Age Group | Hr-HPV Positive Test (% of age group) | Age Group | Hr-HPV Positive (% of age group) |
| 20-29 | 25.1% | 25-29 | 21.1% |
| 30-39 | 16.7% | 30-39 | 11.6% |
| 40-49 | 11.2% | 40-49 | 7.1% |
| 50-59 | 10.5% | >50 | 6.0% |
| 60-69 | 9.7% |  |  |
| 70-79 | 22.0% |  |  |
| >80 | 0.0% |  |  |

When comparing the percentage of HPV types 16 and 18 positive cases between our population and that of the general US female population analyzed from the ATHENA study data by Monsonego et al., HPV type 16 had a lower percentage in our population, 11.7% versus 18.9%, and HPV type 18 had a similar rate in our population, 6.4% versus 7.2%. Our percentages fall very close to that of the general US female public in a study by Dunne et al. [32], 11.7% versus 9.9% for HPV type 16, and 6.4% versus 5.3% for HPV type 18, out of all positive Hr-HPV cases.

## Conclusions

According to our data, Hr-HPV testing shows high-performance measures for detecting high-grade cytology lesions in our women’s veteran population. However, it will miss a significant number of low-grade lesions. Studies have shown that LSIL rarely progresses to CIN3+, primarily when Lr-HPV types cause it. These studies suggest that not detecting a percent of these lesions has minimal to no impact on cervical cancer screening. We believe our findings are in accordance with recent studies and guidelines that recommend primary Hr-HPV testing as the preferred screening method. The total percentage of positive Hr-HPV tests, rates for age, and rates for HPV types 16 and 18 in our women’s veteran population suggest similar HPV prevalence to that of the general US population.

## Data Availability

The source data for this manuscript is confidential and not available to the general public.

